# Prevalence of Chronic Hepatitis B and C in Malaysia- results from a community-based screening campaign

**DOI:** 10.1101/2020.04.30.20085944

**Authors:** ZZ Lim, JS Teo, AC Tan, Tan Soek Siam, Rosmawati Mohamed, KL Goh, Fauziah Jaya, K Senamjit, Azlida Che’ Aun, Rosaida Said, EK Lim, Hamiza Shahar, AH Muhammad Radzi, Tee Hoi Poh, Tan Soon Seng, Jayaram Menon, Rena Menon, TO Lim

## Abstract

**Introduction:** The epidemiology of hepatitis, which is apparently endemic in Asia, is still poorly documented in Malaysia. Available statistics are modelled estimates based on expert input or estimated from small studies on special populations. We therefore determined the prevalence of chronic hepatitis B and C in Malaysia based on a large sample data from a screening campaign.

**Methods:** A total of 10,914 subjects participated in the hepatitis screening campaign in 2018 and 2019. A low-cost Point-of-care test, which has previously been validated, was used to screen for HBsAg and anti-HCV. All screen positive subjects were recalled to undergo confirmatory serology tests and nucleic acid tests.

**Results:** We estimated 1.17% or 238,971 Malaysian adults aged 20 or older had chronic HBV, while only 0.74% or 151,144 adults had chronic HCV. Young adults below age 30 years had very low prevalence of HBV (0.09%). Women had lower prevalence of HBV and HCV, Chinese had the highest prevalence of HBV while Malay had the highest prevalence of HCV.

**Conclusion:** Young adults seems to be protected from HBV perhaps owing to the introduction of universal HBV vaccination since 1989. Chronic HBV however remains prevalent in older adults especially among the Chinese. Chronic HCV is uncommon in Malaysia.

## Introduction

Epidemiological surveillance is poorly developed in many countries where little is known about the diseases prevalent in their populations, and even less is known about the health services provided to address these disease burdens. Such countries are dependent on estimates derived from modelling studies by groups such as the Global Burden of Disease [1,2,3] or studies undertaken on behalf of the World Health Organization [4,5,6]. Likewise in Malaysia despite it being an upper middle income country, we know little about the epidemiology of hepatitis and most other common diseases in Malaysia. Available statistics on hepatitis to date [3–6] are modelled estimates based on expert input or estimated using data from small outdated studies on unrepresentative populations such as from a single institution (eg university), a single convenient group (eg blood donors) or a single at risk group (eg health workers, prisoners or fishermen) [7–22].

A recent study conducted by the Center for Disease Analysis [3] estimated Malaysia has a modelled prevalence of chronic HBV of 0.9% in the period 2005-2011, consistent with the estimate of 0.74% in 1990-2013 from another modelling study funded by the WHO [4]. These prevalence estimates are surprisingly low for Malaysia which is in a region where HBV is endemic [23] but perhaps is consistent with progress made since the introduction of nationwide HBV vaccination program in 1989. Another recent study by the Center for Disease Analysis [5] on behalf of WHO for its Global Hepatitis report [6] estimated Malaysia has a modelled prevalence of chronic HCV of 1.9% in 2011, consistent with the estimate of 2.5% or 450,000 infected people in 2009 with anti-HCV positive from another modelling study [21].

Estimates of disease burden have significant policy consequences. The low modelled prevalence estimate of HBV has led to the termination of antenatal screening and treatment. The high modelled prevalence estimate of HCV was used as the public health justification for the government of Malaysia to issue compulsory license to side step patent on drugs for HCV treatment [24]. Modelled estimate, not being based on any population study, will almost certainly turn out to be wrong, often by a wide margin. For example, Malaysia was estimated to have 3.95 million people infected by Dengue a year in 2010-2013. This was based on a modelling study using the cartographic approach [25]. Subsequent estimate based on actual serial sero-survey data between 2001 and 2013 counted only 2.1 million a year [26].

In this study, we provide estimates of the prevalence of chronic hepatitis B and C in Malaysia based on a large sample data derived from a community-based screening campaign between 2018 and 2019.

## Methods

### Hepatitis screening campaign and Study sample

Screening services for hepatitis are under-developed, and access to anti-viral therapies are limited in Malaysia [27]. In response, a non-governmental organization, the Hepatitis Free Pahang Malaysia (HFPM) [28], has initiated a campaign since 2017 to raise awareness about hepatitis, to provide public screening services and improve access to costly treatments. The campaign is entirely funded by charity and is conducted through partners in each localities or districts in Pahang and other States. We deliberately excluded NGOs or healthcare providers whose members or clientele potentially include persons at high risk of HBV or HCV (such as transgender, sex-worker or former prisoner/drug-user welfare groups, fisherman or other high risk trade associations, HIV/STD or opioid substitution clinics, drug rehabilitation centre or prison hospitals). Study sample for this study are people who attended the above screening campaign in 2018 and 2019. The Ministry of Health’s (MOH) Medical and Research Ethics Committee approved the study and all subjects gave informed consent.

### Registration, screening and confirmatory tests

All participants of the screening campaign are required to register online (see https://hepatitisfreemsia.org.my/page.jsp?pageId=pPatientReg_Disclaimer&type=Screening). The online data system supports the conduct of the screening, manage screen positive subjects for subsequent confirmatory testing and counselling, facilitate reporting of results through short messaging service and helps capture the data for this study.

To screen for HBsAg and anti-HCV, we used a low-cost point-of-care test (POCT, AllTest Biotech Hangzhou China) which we have previously validated [29]. The tests were conducted by trained nurse. The procedure was explained and verbal permission obtained from the participant prior to the testing. Finger-stick capillary samples were taken from participants and the tests were performed according to the manufacturer’s instructions. In the event of invalid result, the test was repeated until a valid result was obtained.

All screen positive subjects were subsequently recalled to undergo confirmatory testing, which were lab based serology tests and nucleic acid tests for HCV RNA and HBV DNA. A trained nurse counselled patients confirmed to have chronic HBV or HCV on infection transmission, risk of liver disease progression, need for monitoring and treatment. Patients with confirmed chronic HBV/HCV were also referred to the local health service for further care. HFPM also funded the direct acting antiviral drugs for some indigent patients with chronic HCV; these were low-cost generic drugs personally imported from India and Bangladesh.

### Statistical methods

The sample size was based on an expected prevalence of 2.0% and precision of the estimate as measured by its 95% exact binomial confidence intervals (CI). For a sample size of 10,000, a prevalence of 2.0% can be estimated with a 95% CI of 1.7 to 2.3, which is deemed sufficiently precise.

Participants in the screening campaign constitute a convenient sample which is not representative of the general population (female, older subjects and Chinese were over-represented compare to the population). To estimate the population prevalence, post stratification [30] was used to adjust the sample totals to known population totals for age, gender and ethnicity based on the Population And Housing Census of Malaysia in 2010. Similarly, prevalence count was estimated by projecting the prevalence estimates to the same 2016 census population projection. We have undertaken a separate validation study of the POCT used in the screening. Using lab based serological test as the diagnostic standard, we determined the POCT for anti-HCV have 98.1% sensitivity and 100% specificity, while that for HBsAg have 95.2% sensitivity and 100% specificity [29]. We use these results to estimate the prevalence of chronic HCV and HBV which are corrected for misclassification due to the use of the POCT screening tests [31].

## Results

A total of 10,912 subjects participated in the hepatitis screening campaign in 2018 and 2019. Table 1 shows the characteristics of all the participants as well as the characteristics of the subjects who were screened positive for HBsAg or anti-HCV. The mean age of the participants was 49 years, there were more female and older subjects, and Chinese were over-represented in the sample. The vast majority of subjects were screened at health fairs organized by local NGO partners and in the state of Pahang.

**Table 1:** Characteristics of subjects who participated in the Hepatitis B and C screening campaign in 2018 to 2019.

|  | All Subjects | HBsAg + | Anti-HCV + |
| --- | --- | --- | --- |
| Total number | 10,912 | 200 | 52 |
| Number Known positive, % | - | 145 | 15 |
| Age, year |  |  |  |
| Mean (SD) | 49(15) | 52(12) | 49(11) |
| Median (IQR) | 50 (37,60) | 51 (42,61) | 50 (40,58) |
| 20-30 years, % | 1416(13) | 3(2) | 3(6) |
| 30-39 years, % | 1877(17) | 36(18) | 10(19) |
| 40-49 years, % | 2182(20) | 57(28) | 13(25) |
| 50-59 years, % | 2610(24) | 46(23) | 15(28) |
| $\geq 60$ years, % | 2827(25) | 58(29) | 11(21) |
| Missing, % | 0(0) | 0(0) | 0(0) |
| Gender |  |  |  |
| Male, % | 4400(40) | 103(51) | 28(54) |
| Female, % | 6512(60) | 97(49) | 24(45) |
| Missing, % | 0(0) | 0(0) | 0(0) |
| Ethnicity |  |  |  |
| Malay, % | 1782(16) | 11(5) | 20(38) |
| Chinese, % | 8519(78) | 183(92) | 30(57) |
| Indian, % | 556(5) | 4(2) | 1(2) |
| Others, % | 55(1) | 2(1) | 1(2) |
| Missing, % | 0(0) | 0(0) | 0(0) |
| Screening method |  |  |  |
| Opportunistic at Clinic/Pharmacy, % | 1111 (10) | 36 (18) | 11 (21) |
| Public event, % | 9801 (90) | 164 (82) | 41 (79) |
| Location of screening by State |  |  |  |
| Pahang | 8715 (80) | 190 (94) | 44 (85) |
| N. Sembilan | 1263 (12) | 7 (4) | 4 (8) |
| Perak | 477 (4) | 2 (1) | 1 (2) |
| Selangor- WP | 383 (4) | 1 (1) | 3 (5) |
| Melaka | 74 (1) | - | - |

We estimated 1.17 percent or 238,971 Malaysian adults aged 20 or older had chronic HBV, while only 0.71 percent or 144,781 had chronic HCV (Table 2).

**Table 2:** Prevalence of Chronic HBV and HCV in Malaysia 2018-2019.

|  | HBV+ |  |  |  | HCV+ |  |  |  |
| --- | --- | --- | --- | --- | --- | --- | --- | --- |
|  | Prevalence rate % | 95% CI |  | Prevalence count | Prevalence rate % | 95% CI |  | Prevalence count |
| All | 1.17 | 1.5 | 0.83 | 238971 | 0.71 | 0.97 | 0.44 | 144781 |
| *By Age* |  |  |  |  |  |  |  |  |
| 20-30 years | 0.09 | 0.28 | 0.03 | 5262 | 0.13 | 0.5 | 0.03 | 7257 |
| 30-40 years | 1.81 | 3.18 | 1.02 | 90931 | 0.58 | 1.24 | 0.27 | 28980 |
| 40-54 years | 1.59 | 2.35 | 1.06 | 84396 | 1.56 | 2.6 | 0.93 | 82721 |
| >=55 years | 1.37 | 2.23 | 0.83 | 58204 | 0.58 | 1.44 | 0.23 | 24625 |
| *By Gender* |  |  |  |  |  |  |  |  |
| Male | 1.49 | 2.18 | 1.02 | 156450 | 0.92 | 1.5 | 0.57 | 96580 |
| Female | 0.82 | 1.24 | 0.54 | 81653 | 0.48 | 0.86 | 0.27 | 47571 |
| *By Ethnicity* |  |  |  |  |  |  |  |  |
| Malay | 0.74 | 1.33 | 0.4 | 71537 | 1.16 | 1.81 | 0.74 | 111625 |
| Chinese | 2.25 | 2.64 | 1.92 | 109241 | 0.37 | 0.55 | 0.24 | 18079 |
| Others | 0.98 | 2.34 | 0.41 | 58136 | 0.23 | 0.95 | 0.06 | 13740 |
| *By Age-Gender* |  |  |  |  |  |  |  |  |
| Male 20-30 years | 0.18 | 0.55 | 0.06 | 5262 | 0.05 | 0.37 | 0.01 | 1494 |
| 30-40 years | 2.12 | 4.62 | 0.96 | 56044 | 0.78 | 2.06 | 0.3 | 20665 |
| 40-54 years | 2.07 | 3.47 | 1.2 | 56342 | 2.41 | 4.38 | 1.3 | 65407 |
| >=55 years | 1.85 | 3.41 | 1 | 38997 | 0.41 | 1.77 | 0.09 | 8719 |
| Female 20-30 years | 0 | 0 | 0 |  | 0.2 | 1.07 | 0.04 | 5733 |
| 30-40 years | 1.46 | 3.17 | 0.66 | 34662 | 0.35 | 1.1 | 0.11 | 8315 |
| 40-54 years | 1.07 | 1.73 | 0.66 | 27830 | 0.67 | 1.63 | 0.28 | 17339 |
| >=55 years | 0.89 | 1.99 | 0.4 | 18983 | 0.74 | 2.36 | 0.23 | 15905 |

Young adults below age 30 years had very low prevalence of HBV (0.09%). Prevalence of HBV and HCV increased with age, and then declined in the oldest subjects (age >=55 years). Men had a much higher prevalence of chronic HBV and HCV than women. Chinese had the highest prevalence of chronic HBV while Malay had the highest prevalence of chronic HCV.

## Discussion

This is one of the largest sero-prevalence study ever conducted in Malaysia; the sample size is even larger than the several sero-prevalence studies on dengue [32], a highly endemic viral infection in Malaysia. The large sample size is necessary to provide precise age-sex and ethnic specific estimates. We found a HBV prevalence of 1.17 percent among Malaysian adults, while the prevalence of HCV was only 0.74 percent. The age trend in HBV prevalence suggests young adults age below 30 years were protected from HBV perhaps owing to the introduction of universal HBV vaccination since 1989, while lower prevalence in the oldest age group is likely due to pre-mature mortality from progression to cirrhosis and hepatocellular carcinoma [23]. Male had higher prevalence for both HBV and HCV, as expected. Similarly, Chinese had higher prevalence of HBV, while Malay had higher prevalence of HCV.

HBV has long been endemic in the Asia-Pacific region, though this has declined in many countries since the advent of universal vaccination in the 1990s [23]. Table 3 summarizes recent prevalence estimates in adults derived from large population based studies from some countries in the region. Our estimate is consistent with the observed decline in recent HBV sero-prevalence in several countries except Mongolia in the region.

**Table 3:**
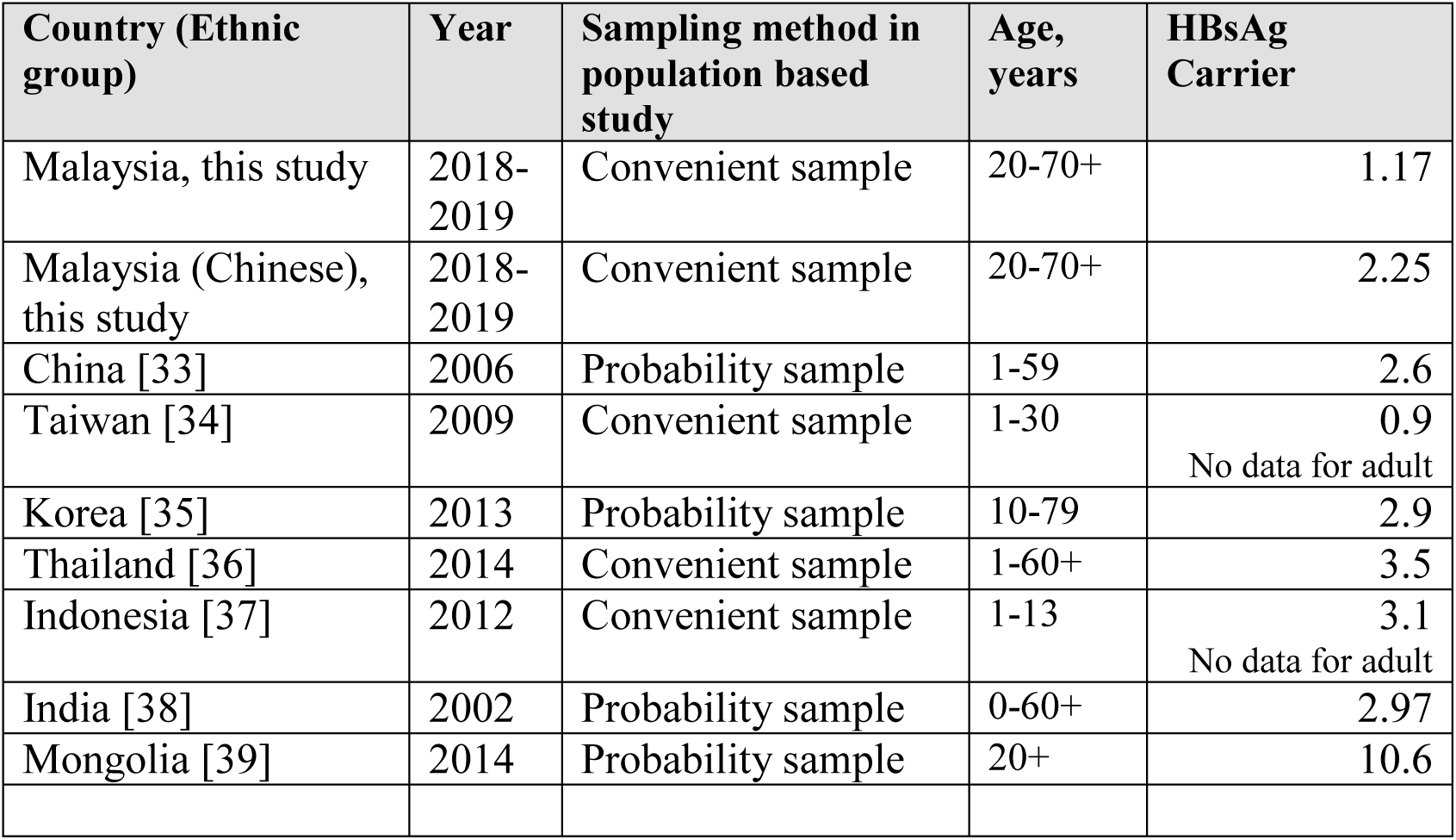
Recent sero-prevalence of HBV from population based studies in some Asia-Pacific countries.

HCV is highly endemic in Central Asia and Mediterranean but not in the Asia-Pacific region [23]. Unlike for HBV, few recent population based sero-prevalence surveys have been conducted. We summarize the prevalence estimates in Table 4. Our low estimate of HCV prevalence is consistent with those observed in China, Thailand and India, though like for HBV, Mongolia has exceptionally HCV prevalence in the region.

**Table 4:**
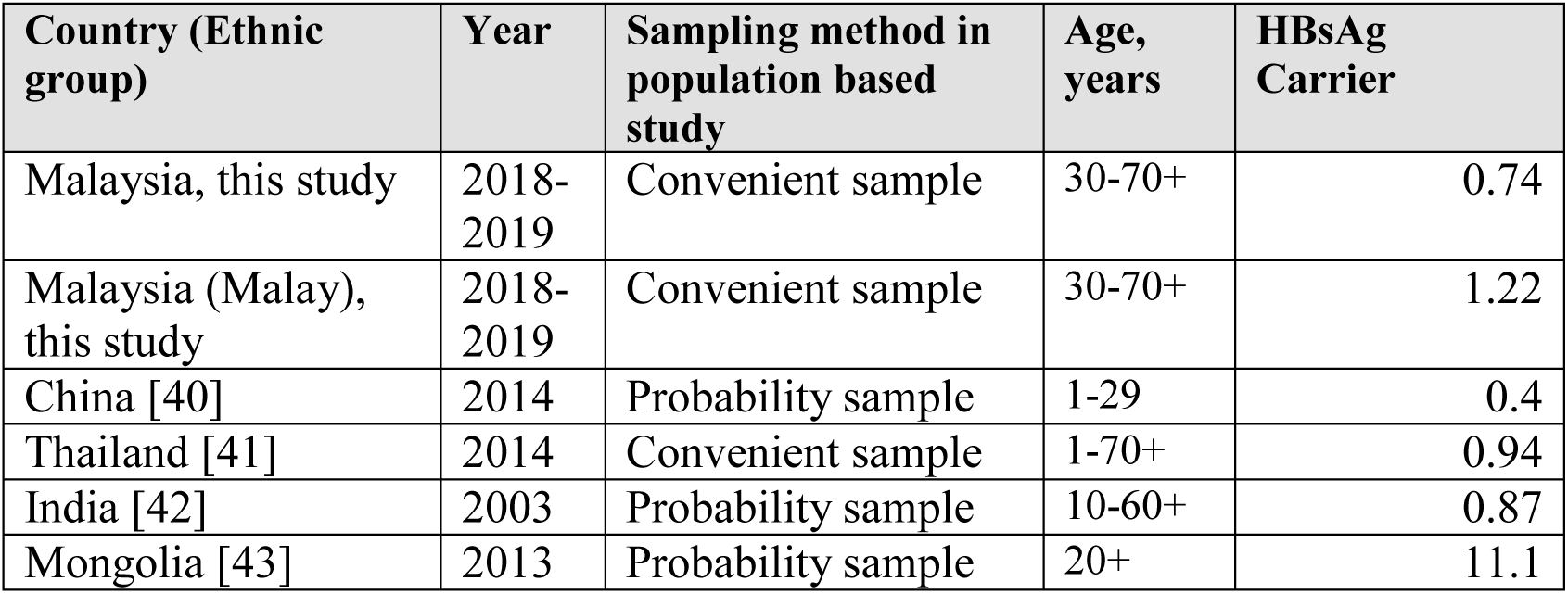
Recent sero-prevalence of HCV from population based studies in some Asia-Pacific countries.

| Country (Ethnic group) | Year | Sampling method in population based study | Age, years | HBsAg Carrier |
| --- | --- | --- | --- | --- |
| Malaysia, this study | 2018-2019 | Convenient sample | 30-70+ | 0.74 |
| Malaysia (Malay), this study | 2018-2019 | Convenient sample | 30-70+ | 1.22 |
| China [40] | 2014 | Probability sample | 1-29 | 0.4 |
| Thailand [41] | 2014 | Convenient sample | 1-70+ | 0.94 |
| India [42] | 2003 | Probability sample | 10-60+ | 0.87 |
| Mongolia [43] | 2013 | Probability sample | 20+ | 11.1 |

Our study has several limitations. First, the study subjects were not a probability sample and is not representative of the population. Hence, there were more female and older subjects than in the general population, Chinese and Pahang residents were over-represented as a result of the conduct of the campaign through local NGOs, most of which were local faith or ethnic based organizations in Pahang. Post-stratification was required to adjust the sampling weight to reflect the age, sex and ethnicity distribution of Malaysia. Second, subjects known to have hepatitis may be more or less willing to participate in screening. This source of bias applies to probability sample too; such subject may be more or less willing to consent to be tested in a probability sampling survey. The risk of this bias however is lessened in our study by pooling the data from numerous (109) screening events or venues conducted in numerous rural and urban locations spread over a wide geographical region. Third, for operational and cost reasons, we have used a POCT for screening instead of lab based serology tests. The POCT has been validated, and the prevalence estimates reported here are corrected for the misclassification bias due to the use of POCT.

## Data Availability

The datasets used and/or analysed during the current study are available from the corresponding author on reasonable request

**List of abbreviations**
CI: Confidence interval
HBV: Hepatitis B virus
HCV: Hepatitis C virus
HFPM: Hepatitis Free Pahang Malaysia
MOH: Ministry of Health Malaysia
NGO: Non-Governmental Organizations
POCT: Point of Care Tests
SD: Standard deviation
WHO: World Health Organization

## Declarations

### Ethics approval and Consent

The Ministry of Health’s (MOH) Medical and Research Ethics Committee approved the study. All subjects who participated in the screening campaign gave written informed consent

### Consent for Publication

Not applicable. The manuscript does not report on any individual participant’s data in any form (images, videos, voice recordings etc).

### Author’s contributions

TSS, RM, GKL, FJ, SK, ACA, RS, LEK, HS, MR, THP, TSS, RM contributed to the subject matter expertise. They also contributed to the writing of the manuscript. LTO, LZZ, TJS, TAC conceived the idea behind this study and contributed to the study design, survey conduct, data analysis and interpretation, report writing and subject matter expertise. All authors read and approved the final manuscript.

## Acknowledgement

The authors would like to extend their sincere gratitude and appreciation to all the staff and volunteers of Hepatitis Free Pahang Malaysia for their efforts in conducting the screening campaign. We also wish to thank all those whose names are not mentioned here who render their excellent service especially during the data collection.

